# Reliable machine learning models in genomic medicine using conformal prediction

**DOI:** 10.1101/2024.09.09.24312995

**Authors:** Christina Papangelou, Konstantinos Kyriakidis, Pantelis Natsiavas, Ioanna Chouvarda, Andigoni Malousi

## Abstract

Machine learning and genomic medicine are the mainstays of research in delivering personalized healthcare services for disease diagnosis, risk stratification, tailored treatment, and prediction of adverse effects. However, potential prediction errors in healthcare services can have life-threatening impact, raising reasonable skepticism about whether these applications are beneficial in real-world clinical practices. Conformal prediction is a versatile method that mitigates the risks of singleton predictions by estimating the uncertainty of a predictive model. In this study, we investigate potential applications of conformalized models in genomic medicine and discuss the challenges towards bridging genomic medicine applications with clinical practice. We also demonstrate the impact of a binary transductive model and a regression-based inductive model in predicting drug response and the performance of a multi-class inductive predictor in addressing distribution shifts in molecular subtyping. The main conclusion is that as machine learning and genomic medicine are increasingly infiltrating healthcare services, conformal prediction has the potential to overcome the safety limitations of current methods and could be effectively integrated into uncertainty-informed applications within clinical environments.

## 1 Introduction

AI-based models are having a transformative impact on high-risk predictions made for personalized medicine applications.^1^ Genomic medicine as a cornerstone of precision medicine has the potential to revolutionize healthcare for rare diseases and cancer through robust and reliable personalized diagnosis, risk stratification, and tailored treatment solutions. ^2^ However, prediction errors can have life-threatening impact, raising reasonable skepticism on whether these applications are beneficial in routine clinical practices. The main sources of prediction errors are the stochasticity and complexity of the models, the different data collection/curation protocols, and domain shifts that result in data falling outside training distributions. ^22^

### 1.1 Uncertainty quantification methods

To mitigate the risks of singleton predictions in zero-tolerance healthcare applications, several methods calibrate the outcomes into distribution predictions made for each sample. ^20^ Bayesian techniques like Monte Carlo dropout, ^37^ variational inference,^45^ and non-Bayesian methods such as deep ensemble, ^73^ softmax calibration, and selective classification^74^ rely on prior distributions and posterior inference to provide a probabilistic framework for estimating uncertainty in predictions. These approaches estimate a probabilistic distribution instead of deterministic outcomes, enhancing decision-making and model interpretability, which is particularly valuable in healthcare settings for decision support and risk mitigation. Additionally, distribution-free uncertainty quantification techniques offer a general framework with rigorous statistical guarantees to black-box models, reducing uncertainty in decision-making processes.

In this context, Conformal Prediction (CP) stands out as an effective and versatile method for quantifying statistically rigorous uncertainty (Angelopoulos et.al, 2021). Unlike traditional prediction methods, CP generates prediction sets with guaranteed error rates rather than point estimates and operates under the assumption of independent and identically distributed random variables (i.i.d.), emphasizing exchangeability.

### 1.2 Principles of conformal prediction

CP was initially proposed by Vladimir Vovk (2005),^11^ and later expanded by Vovk and Shafer in 2008.^12^ CP provides a formal and structured approach to addressing questions that were often approached in a vague and abstract way in machine learning (ML) models, such as determining the confidence level of a model’s predictions. CP quantifies these uncertainties by estimating prediction intervals for regression problems and a set of classes for classification problems. Both prediction intervals and classes are guaranteed to include the actual value with a predefined confidence level. Practically, CP estimates how “unusual” a sample seems to be relative to previous ones. Therefore, CP uses past experience to determine accurate confidence levels in new predictions.^12^ The prediction areas are set by including samples that have very common values or better yet, those that are not very unlikely. CP operates under the assumption of “independent and identically distributed random variables (i.i.d.)”, emphasizing the exchangeability assumption. This assumption implies that the order of observations does not affect their joint distribution. This makes CP particularly valuable in real-world biomedical applications in which making assumptions about the underlying data distribution may be challenging or unrealistic.

### 1.3 Conformal prediction frameworks

CP is defined as a mathematical framework that can be used with any ML model to produce reliable predictions with high probability and user-defined error rates. ^11,12^ Given a set of training data *D*, with *n* instances *{*(*x_i_, y_i_*), …, (*x_n_, y_n_*)*}*, where *x_i_* is a feature vector and *y_i_* is the true label of the *i*-th sample, with labels in *Y* = [1*, K*], the objective is to predict the label *y ∈ Y* for a new sample with feature vector *x_n_*_+1_. In classification problems, we test all possible classes of a new instance and measure the probability of a prediction to be the correct one for each class. To do so, we calculate the *non-conformity* score *α_i_*, which is based on the underlying ML algorithm and indicates how strange an instance is compared with other instances. A simple example of a *non-conformity* score is the *1-predicted probability of the true class*, otherwise called inverse probability. Based on the hypothesis that the instances are independently and identically distributed random variables (i.i.d.), for a new instance *x_n_*_+1_ we compute the *non-conformity* score 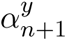 for each possible class. Finally, for each possible label we calculate the *p-value* as:

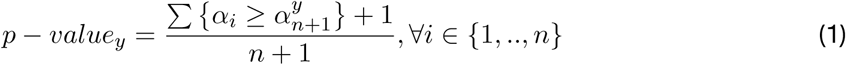

*p-value* is used to evaluate the *non-conformity* score of the new instance *α_n_*_+1_ against all other *non-conformity* scores. False predictions result in a higher *α_n_*_+1_ than the rest *non-conformity* scores of the training set. In this case, we get a low *p-value*, while in cases of correct prediction, we expect a higher *p-value*. So, for a dataset that satisfies the i.i.d. assumption, every *p-value* in Eq. 1 has the following property validity guarantee:

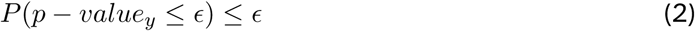

where, *ϵ* is the user-defined significance level (or target probability error). This statement, *P* (*p− value_y_ ≤ϵ*), expresses the probability P that the *p-value*, derived from a set of independent and identically distributed (i.i.d.) instances, falls below or equals the user-defined significance level *ϵ*. This probability is constrained by the property in Eq. 2, signifying that the likelihood of obtaining a *p-value* less than or equal to *ϵ* is itself limited by *ϵ*. In practical terms, this encapsulates the assurance that, under the assumption of i.i.d. instances, the probability of observing a *p-value* leading to the rejection of the null hypothesis does not exceed the chosen significance level. Consequently, we may output a set of possible predictions and construct the prediction region *C^ϵ^*, as follows:

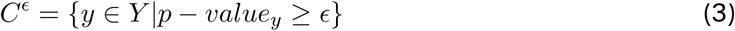

Because of the property in Eq. 2, the probability that each set of predictions does not contain the correct class will be less than or equal to *ϵ*, so we limit the error rate to less than or equal to *ϵ*. In a binary classification problem with a positive and a negative class there are four possible outcomes for a conformal prediction i.e., positive, negative, both classes (positive and negative), and no class assignment (empty class). In each case, the classes are included in the prediction region when we are confident with the desired level. The “empty” label indicates that the sample lies outside the range where the model can make reliable predictions. In other words, the model cannot assign any class with the user-defined required confidence level, signifying that the sample is beyond the boundaries of the model’s applicability. Consequently, the classification decision needs to be determined, by other in silico methods and subsequently integrated into an enriched model. This step is useful for expanding the model’s applicability domain. ^9^

In a regression analysis framework, CP transforms point predictions from a model 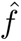 trained on *D* data with *n* instances, to intervals which contain the true value with a level of guarantee defined by the user. In this case, to compute the *non-conformity* scores for every sample in the training set, we measure how different the observed *y_i_* is from the model prediction 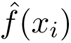. A simple measure to calculate non-conformity is the absolute residual: 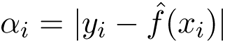. Given *ϵ* the user-defined significance level, we calculate the *Q*_1_*_−ϵ_* quantile of the scores as:

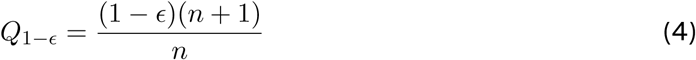

For a new input *x_n_*_+1_, the prediction interval is defined as follows:

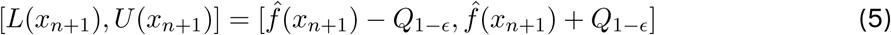

where, L is the lower limit and U the upper limit for the new input *x_n_*_+1_. The resulted prediction interval [*L*(*x_n_*_+1_)*, U* (*x_n_*_+1_)], assuming data *D* are exchangeable, satisfies the property of marginal coverage:

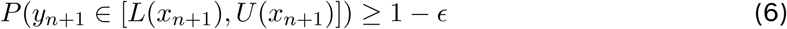

In other words, the probability that the predicted value is included in the prediction interval is bigger or equal to the user-defined level of confidence.

#### 1.3.1 Transductive conformal prediction

CP was originally used in the transductive or full version. ^11,13^ The transductive CP (TCP) uses all the available data to train the model and thus, we can produce more accurate and informative predictions. After choosing the appropriate *non-conformity* function, we add the features of a new instance *x_n_*_+1_, and assuming its class *y_n_*_+1_, we retrain the model K times, where K is the number of all the possible classes for *x_n_*_+1_ (Fig.1). In a binary classification problem, the model will be trained 2*Z times for each class, with Z being the number of points in the test set. Then, for these two new training sets, we apply the *non-conformity* measure, we compute the *p-values* (Eq. 1) and finally, we check whether the features (*x_n_*_+1_) of an instance in the test set “conforms” to the predictions of the training set and leads to decisions for the creation of the prediction region. TCP is a suitable method for analyzing small data sets as it works as an online framework. For larger datasets more computationally efficient methods should be selected e.g., inductive CP.

**Figure 1:**
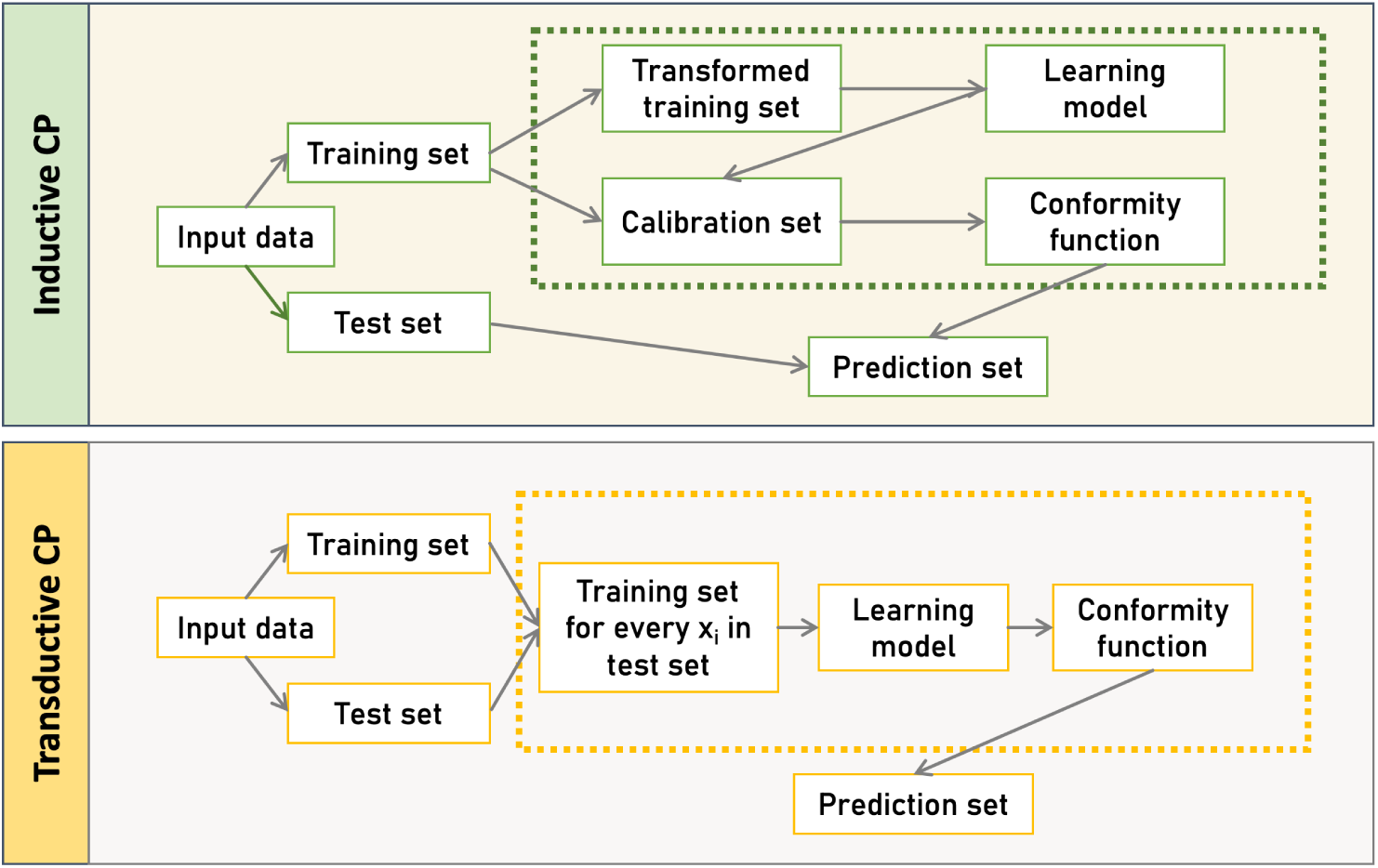
CP-based frameworks. Inductive CP (ICP), involves a static split of the dataset into training and calibration sets, using a single model for all predictions, while Transductive CP (TCP), builds a new model for each test instance, offering potentially more tailored predictions at the cost of increased computational effort.

#### 1.3.2 Inductive conformal prediction

Inductive CP (ICP) is the most popular CP approach. TCP has high a computational cost and may not be suitable for certain applications in genomic medicine. For example, multi-omics analyses usually involve large datasets due to the sheer size, complexity, and variability of the genomic data and the technologies that are used to produce it. To deal with this issue ICP trains the basic algorithm only once^14^ by splitting the training set n into two smaller sets, a training set with *m < n* and a *calibration set* with *n− m* instances. The training set is used to create the “prediction region” and the instances in the *calibration set* are exclusively used to calculate the *p-value* of each possible class of a new test instance *X* (Fig. 1). No matter which CP method we use, ICP will result in unbiased predictions. The efficiency of an ICP model depends on many factors such as, how large and well-constructed (i.i.d.) the dataset is, how effective is the underlying ML algorithm, and which *non-conformity* measure is employed.

### 1.4 Evaluation and parameterization of CP methods

Conformal predictors enhance the reliability of black-box models by generating prediction sets that reflect uncertainty in high-risk applications. The evaluation of the conformalized model generally concerns adaptivity, size, and coverage of the prediction intervals. As Angelopoulos et al. (2021) proposed, a model’s adaptivity can be assessed by the size of the prediction sets, with larger sets indicating higher uncertainty and more challenging predictions, while smaller sets signify easier ones (Angelopoulos et.al, 2021). Adaptivity is closely linked to the model’s conditional coverage, which ensures that the true label falls within the prediction region at a defined confidence level for any subset of the test set. Marginal coverage is achievable, but conditional coverage requires consistency across subsets of the test data. Angelopoulos et al. recommended the size-stratified coverage (SSC) to measure adaptivity and suggested verifying conditional coverage by repeating the framework with different combinations of calibration and test sets (Angelopoulos et.al, 2021). In a similar vein, Park et al. proposed the meta-XB, a meta-learning approach designed for cross-validation-based CP that focuses on reducing the average size of prediction sets while ensuring formal calibration for each task. ^47^

Given that CP can be applied across various prediction models, its effectiveness hinges on three critical parameters: the choice of the non-conformity function, the size of the calibration set, within an ICP, and the underlying model.

#### 1.4.1 Non-conformity function

The choice of the *non-conformity* function is critical for the effectiveness of conformal predictors. This function evaluates how “strange” or dissimilar a new instance is relative to the training data. Vovk et al. introduced this measure to gauge dissimilarity, although it can also be adapted to a conformity measure for assessing similarity. ^11^ Non-conformity measures are derived through the application of a *non-conformity* function, and the selection of this function depends on the nature of the underlying model. While any function can theoretically serve as a *non-conformity* measure, choosing the most appropriate one can significantly impact the efficiency and accuracy of a conformalized model.

#### 1.4.2 Calibration set size

Another crucial parameter in conformal prediction is the size of the calibration set. This size affects the amount of data used to initially train the model, with a larger calibration set potentially improving model coverage (Angelopoulos et.al, 2021). However, dealing with large datasets can increase computational costs. To address this challenge, several methods have been proposed. Abda et al. scaled the TCP framework on large real-world data, reducing computational cost while maintaining CP guarantees (Abad et.al, 2022). Schuster et al. introduced CATs, an extension of CP designed to accelerate inference in transformer models for natural language processing applications while ensuring high confidence (Schuster et.al, 2021). Additionally, a dynamic CP framework trains models with iteratively updated samples.^48^ Variations in CP conceptualization include regularization techniques, ensemble methods, and graph-based approaches to minimize data dimensions (Davis et.al, 2024). ^49–51^

#### 1.4.3 Underlying model selection

The effectiveness of conformal prediction (CP) is closely tied to the choice of the underlying learning model. The non-conformity measure, which assesses how unusual a new instance is relative to the training data, depends heavily on the model’s performance. Different models may exhibit varying degrees of effectiveness and reliability within a CP framework. Therefore, selecting an appropriate model is crucial for generating accurate and reliable prediction intervals. The performance of the non-conformity function is influenced by how well it aligns with the model’s predictive capabilities. A well-calibrated and accurate model will produce better-calibrated conformal intervals. As a result, optimizing the model’s parameters and ensuring its suitability for the specific application are essential steps before implementing and evaluating conformal prediction.

### 1.5 Conformal prediction and distribution shift

A major concern in ML applications is the deviation of the properties and distribution of the new, unseen data compared to those of the training set. The so-called distribution shift is frequently observed in real-world predictive models, when the joint distribution of inputs and outputs differs between training and test stages. Covariate shift occurs when there is a discrepancy between the distributions of input points in the training and the test datasets, even though the conditional distribution of output values given input points remains consistent. ^41^ The weighted CP proposed by Tibshirani et al. ^38^ can handle covariate shift by weighting each *non-conformity* score by a probability that is proportional to the likelihood ratio of the new data distribution to those used to build the model. The maximum mean discrepancy (MMD), proposed by Borgwardt et al., ^40^ is a kernel-based statistical test used to determine whether two samples are drawn from different distributions. In contrast to typical measures like Kolomogorov-Smirnov test that can only be applied in vectors, MMD is applicable in multivariate data that are frequently met in genomic data analyses. The null hypothesis in MMD statistical test states that there is no difference between the distributions of the two datasets and therefore that the datasets are drawn from the same distribution.

The label or prior probability shift, refers to a shift in the distribution of class variables. A variation of CP named Mondrian CP (MCP) can remedy this difference between the train and validation samples. In MCP, each class is evaluated independently to determine the confidence of assigning an instance to that class. Predictions for the calibration set produce *non-conformity* scores for each class. MCP ensures controlled error rates by categorizing training sets based on features or their combinations and defining significance for each category. ^11,54^ It compares non-conformity scores only within the same category, making it suitable for poorly distributed datasets. Label Conditional Mondrian Conformal Prediction (LCMCP) is a specific case of MCP where the category of each instance is determined by its label. Under the same scope, Bostrom et al. proposed the Mondrian conformal regressors handling the range of the prediction interval. ^53^ Sun et al. proposed a Mondrian Cross-Conformal Prediction for large imbalanced bioactivity datasets, and proved that this framework performs well in this type of shifts.^52^

Recent work suggests CP as an effective framework that can handle distribution shifts. Cai et al. utilized an Inductive Conformal Anomaly Detection (ICAD) approach for online detection of distribution shifts on high-dimensional data with low computational cost and efficiency. ^55^ Hernandez et al. demonstrated the robustness of conformalized models in predicting the activities of novel molecules on cancer cell lines, offering valuable insights for drug discovery under strong distribution shifts (Hernandez et.al, 2024). However, in real world applications, distribution shifts are commonly encountered with unexpected results in model performance. For large scale datasets, black-box model architectures or hidden distribution shifts, predictions must undergo careful examination before being applied in clinical decision making. To prove this Kasa et al. examined how those shifts affect CP and concluded that the performance degrades and the coverage guarantees are frequently violated, highlighting the challenges and the need for further elaboration on these issues (Kasa et.al, 2023).

## 2 Conformal modelling applications in biomedicine

### 2.1 Applications in biomedical imaging

In principle, CP coupled with any traditional learning model can be used to address uncertainty in a wide range of scientific domains. In medical applications, it is crucial for any predictive model to generate predictions tailored to each individual patient rather than relying on generalizations from a broader population. Hence the definition of the confidence intervals for individual predictions is critical especially when these models are adopted in clinical environments. ^3^ In such clinical applications CP is used to intuitively express the uncertainty of a prediction and to facilitate the model’s transparency and robustness (Lu et.al, 2022). For example, CP has been employed in medical imaging applications for subgroup analysis, distribution shift estimation, and for the elimination of prediction errors in safety-critical applications.^23,61^ Using microscopic biopsy images Olsson et al. implemented an effective CP-based model for diagnosis and grading of prostate cancer. ^4^ Additionally, Kapuria et al. proved that, using CP, clinicians can make informed decisions and minimize the risk of colorectal cancer polyps misdiagnosis.^62^ In the same context, Lambrou et al., applied a CP approach coupled with Genetic Algorithms to diagnose breast cancer based on digitized images of fine needle aspirates from breast masses.^10^ In non-cancer applications, CP was used by Lu et al. to develop a deep learning model for grading the severity of spinal stenosis in lumbar spine MRI^24^ and Wieslander et al. combined deep learning methods with CP to predict tissue sub-regions using hierarchical identification on rat lung slides.^25^

### 2.2 Applications in drug discovery

In preclinical settings, CP has been applied in drug discovery, mainly to predict the biological activity of compounds based on their chemical structure. CP-based methods have been used as an alternative approach to traditional QSAR models, to predict target-ligand binding that are enriched with uncertainty estimates (Xu et.al, 2023). ^26^ For example, Alvarsson et al. used CP on top of random forest models to classify three different ATP transporters. ^9^ The authors concluded that the higher the level of confidence the larger the prediction interval or set of predictions, and they suggested CP as an effective method for drug discovery applications. Toccaceli et al. demonstrated the application of an Inductive Mondrian Conformal Predictor to predict the biological activities of chemical compounds by addressing challenges such as the large number of compounds, the high dimensionality of the feature space, the sparseness, and the class imbalance.^6^ In the same context, CPSign proposed a conformal predictor that is applied to chemical descriptors for chemoinformatics modeling (McShane et.al, 2023) while several other applications in biomolecular design proposed sophisticated methods to handle covariate shift, enabling the computation of distribution-free prediction intervals. ^27,60^ Similar CP approaches have been extensively applied in modeling chemical compound toxicity. ^5,7,8,28,29^

## 3 Genomic medicine and conformal prediction

Despite their widely recognized contribution to medical imaging and drug discovery, conformal predictors have not been sufficiently used in joint applications of genomics and medicine. Genomic medicine is an emerging medical discipline and a rapidly evolving field of predictive modeling applications. In areas such as oncology, pharmacology, rare or undiagnosed diseases, and infectious diseases genomic medicine has a transformative impact on improving medical decisions, and advancing medical knowledge, and healthcare delivery.

### 3.1 Dealing with uncertainties of genomic medicine models

To advance clinical applications, genomic medicine models must deal with a variety of uncertainty-inducing and safety-critical issues that are mainly caused by the inherent complexity and variability of the biological systems, the inter-individual heterogeneity in genetic profiles, environmental exposure, and lifestyle as well as the non-linearity of the interactions within the patients data. Uncertainty manifests in various steps of genomic analysis, and particularly for ML applications has different dimensions. Uncertainties might involve the ambiguity, complexity, or deficiency of the data, as well as the unpredictability of the models. It is important to understand the dimensions of uncertainty, however, it is also important to recognize that uncertainty is not always problematic. ^30^ Uncertainty estimates can help acknowledge the complexity of molecular events and account for the data variability in a model recalibration. By definition, CP estimates uncertainty when making personalized decisions and leverages the evidence linking each in-dividual’s genetic makeup to zero-tolerance applications such as medical decision-making, diagnosis, risk assessment, and treatment strategies. In this context, CP-enriched models can greatly benefit from the availability of massive amounts of trainable multi-omics data derived from high-throughput sequencing technologies and they can in turn contribute to improved generalizability and calibration of rare events of the learning models.

### 3.2 Current landscape of conformal prediction

In the field of genomic medicine only few CP uncertainty-aware models have been reported in the literature. Ianevski et al. used patient-derived single-cell transcriptomic data to train a gradient boosting model that prioritizes multi-targeting therapeutic compounds for stratified cancer treatment (Ianevski et.al, 2023). In this ex vivo drug testing methodology the conformalized model was built using subclone-specific differentially expressed genes and helped to filter out predictions with low conformity scores. Single-cell transcriptomic data was also used by Sun et al. to identify subtypes within the neural stem cell lineage.^31^ In this work, CP is part of a general framework for estimating uncertainty in spatial gene expression predictions and is applied to calculate the calibration score that links the cell-centric variability to the prediction error.

In a different setting, Sun et al. proposed a method to address personalized genetic risk assessment for complex diseases that relies on a Mondrian cross-conformal prediction model to estimate the confidence bounds of the polygenic risk score prediction. ^32^ The proposed method showed that using the predicted risk of each individual to classify as a case or control is more clinically relevant than group-wise assignments to high-risk or low-risk groups based on an arbitrary selection of the extreme scoring samples.

On the protein level, conformal predictions have recently been employed as an effective approach to detect protein homologies enabling the discovery of new proteins with likely desirable functional properties (Boger et.al, 2024). The method provides statistical guarantees of the homology searches of a query protein against a lookup database -instead of protein pairs- and functional annotations by leveraging the vast amount of protein structures produced by algorithms such as Alphafold. ^63^ The proposed conformalized protein mining method has potentially significant implications in genomic medicine including drug repurposing utilizing proteins with unique and desirable features, the development of therapeutic enzymes or monoclonal antibod-ies for personalized disease treatment and engineering proteins for enhanced stability, activity, or binding affinity, creating more effective therapeutics.

In pharmacogenomics, prediction error estimates have been employed in a CP model to predict drug sensitivity and prioritize drugs using gene expression levels of cancer cell lines.^64^ The prediction outcomes show substantial improvement of CP prediction accuracy and highlight the importance of developing more sophisticated methods that incorporate multi-omics data, to address not only monotherapies but also combinatorial drug delivery.

### 3.3 Perspectives in the era of precision medicine

CP can be an essential component for a much wider range of genomic medicine applications combining predictive modelling and high-risk decision-making. Genomic ML applications with clinical relevance can greatly benefit by uncertainty estimates in the following fields:

#### 3.3.1 Variant calling and prioritization

The diagnosis and disease risk assessment in genomic medicine is most often based on the presence of genetic variants. In next-generation sequencing studies, genetic variants are detected by complex deep neural network architectures, e.g. DeepVariant ^42^ and DeepSNV.^43^ However, accurate variant calling is not a straightforward process and is often error-prone, especially for tumor samples with high heterogeneity and low purity, or for genomic regions that are difficult to map. ^33^ To be able not to take the risk of a prediction could be of great clinical significance, particularly while trying to distinguish between somatic and germline variants or to prioritize rare variants. In addition to variant calling, prioritizing the detected variants based on their functional effect introduces challenges that can be of clinical relevance when sorting neutral or deleterious variants among those of unknown significance.

#### 3.3.2 Immunotherapy response prediction

In a similar setting, the mutational load of tumor DNA samples, known as tumor mutational burden, is a strong predictor of response to immunotherapy. However, several issues, including the variability of response levels by cancer type and the lack of a standardized method for calculating variant burden, limit the reproducibility and reliability of the predictions. In this context, conformalized learning models are suitable for estimating the uncertainty of the immunotherapy response predictions, and to avoid to take the risk of a prediction in inconclusive cases.

#### 3.3.3 Pharmacogenomics

Besides predicting immunotherapy responses, the genetic makeup is a mainstay of research in pharmacogenomics to tailor therapeutic solutions either by identifying biomarkers of pharmacological response or by developing learning models. ML-based applications develop strategies to prioritize candidate anti-cancer drug compounds, or predict the sensitivity levels of a particular compound, yet out of the context of reliability testing and uncertainty estimates.^34,35^ Recently, Lenhof et al. developed a conformalized approach that predicts and prioritizes drug sensitivity on cell line-based monotherapy responses, based on gene expression profiles and user-defined certainty levels. ^36^ Compared to cell lines, patient-derived profiles are preferred in the development of clinical pharmacogenomic models however, they introduce additional complexities that increase the uncertainty and risk of an erroneous prediction. In addition, novel approaches demonstrate the need to integrate multi-omics data in drug response predictions, including mutations, copy number variations and proteomics. Multi-omics data can be particularly informative and, when combined with uncertainty estimates, could facilitate safer predictions and decipher the physical/functional gene-drug interactions. These potential applications collectively demonstrate the need to establish robust genomic medicine frameworks capable of evaluating the predictability in clinical applications and enhancing reproducibility.

#### 3.3.4 Reverse vaccinology

Reverse vaccinology (RV) is a rapidly evolving approach in vaccine development against pathogens that utilizes genome sequences to predict antigens that can elicit strong immune responses. RV workflows include several analysis steps (Trygoniaris et.al, 2024) in which ML models are often used to predict B-cell and T-cell epitopes based on the pathogen’s genomic and proteomic features. ^69,70^ In addition, predictive models are used to assess and prioritize vaccine candidates based on factors like antigenicity, immunogenicity, conservation across strains, and homology to host proteins to avoid autoimmune reactions. ^71^ A critical step in RV is the integration of 3D modelling algorithms to predict the folded 3D structure of the vaccine construct and the development of multi-epitope vaccines. Considering the poor quality of the training data and the difficulties in experimental screening, being able to quantify uncertainties in each ML-based analysis would greatly advance model calibration and validation. ^72^ In this context, CP models can be particularly useful in validating ML predictions by ensuring that the specified coverage probability is maintained across different datasets and pathogen-host application scenarios.

#### 3.3.5 Antimicrobial resistance

Antimicrobial resistance (AMR) is a serious public health threat that is responsible for prolonged hospitalizations and more than one million deaths per year. ^65^ The availability of millions of whole genome sequencing data annotated with diverse AMR phenotypes enabled the development of ML methods that predict AMR using pathogens features, mainly genomic variability ^66,67^ and biochemical information. ^68^ However, the reliability of the predictions is subjected to several confounding factors e.g., biased sampling and poor genome assembly quality due to increased contamination rates, poor coverage and low read depth. Erroneous predictions of AMR against antibiotic compounds can be life-threatening and therefore uncertainty guarantees in either supervised classification (sensitivity/resistance prediction) or regression problems (quantification of the minimum inhibitory concentration values) can be particularly valuable. In this context, conformalized models can be important preventive measures offering safer clinical decision making, while also helping in deciphering the molecular mechanisms underlying AMR.

In this study, we rigorously explore the potential of conformal predictors in genomic medicine and demonstrate their pivotal role in yielding more reliable predictions using three application scenarios. Specifically, we evaluated CP-enriched models on a binary classification, on a multiclass classification problem under distribution shift and a regression-based application aiming to gain further insights into how conformalized predictive modeling can be practically integrated into genomic medicine. The study discusses further the strengths and challenges and highlights the main issues that should be addressed in order to unequivocally ensure patient safety when pivotal decisions are delegated to clinically deployed AI systems.

## 4 Experimental setting and results

To practically assess the applicability of CP in genomic medicine we sought to examine how ML models can benefit from conformalized predictions in two exemplar classification and in one regression problems. The objective was to cover both binary and multi-class predictions, small and larger datasets, different application domains and both inductive and transductive frameworks. First, a TCP-based pharmacogenomic learning model was implemented to demonstrate the impact of conformal predictors in tailoring personalized therapeutic decisions. Transcriptomic profiles of rheumatoid arthritis and Crohn’s disease patients undergoing infliximab treatment were used to estimate the uncertainty of the drug sensitivity predictions (Fig. 2). In the multi-class setting, an inductive conformal predictor was built to assess the diagnostic predictions for patients with different transcriptional subtypes of diffuse large B-cell lymphomas (Fig. 2). Finally, in the regression setting, an inductive conformal predictor was used to predict the pharmacological response of cancer cell lines to afatinib. Both classification models used publicly available gene expression datasets deposited in Gene Expression Omnibus (GE under the accession IDs GSE42296 for rheumatoid arthritis and Crohn’s disease^15^ and GSE181063 for diffuse large B-cell lymphoma samples. To train the regression model, data from the Genomics of Drug Sensitivity in Cancer (GDSC) database was used. In all use cases, we applied MRMR (Maximum Relevance - Minimum Redundancy) feature selection method and statistical tests to assess the validity of the i.i.d. assumption. ^46^ It should be noted that although the application scenarios address real-world research problems, the prediction results are not intended to produce novel research findings as this is out of the scope of this review.

**Figure 2:**
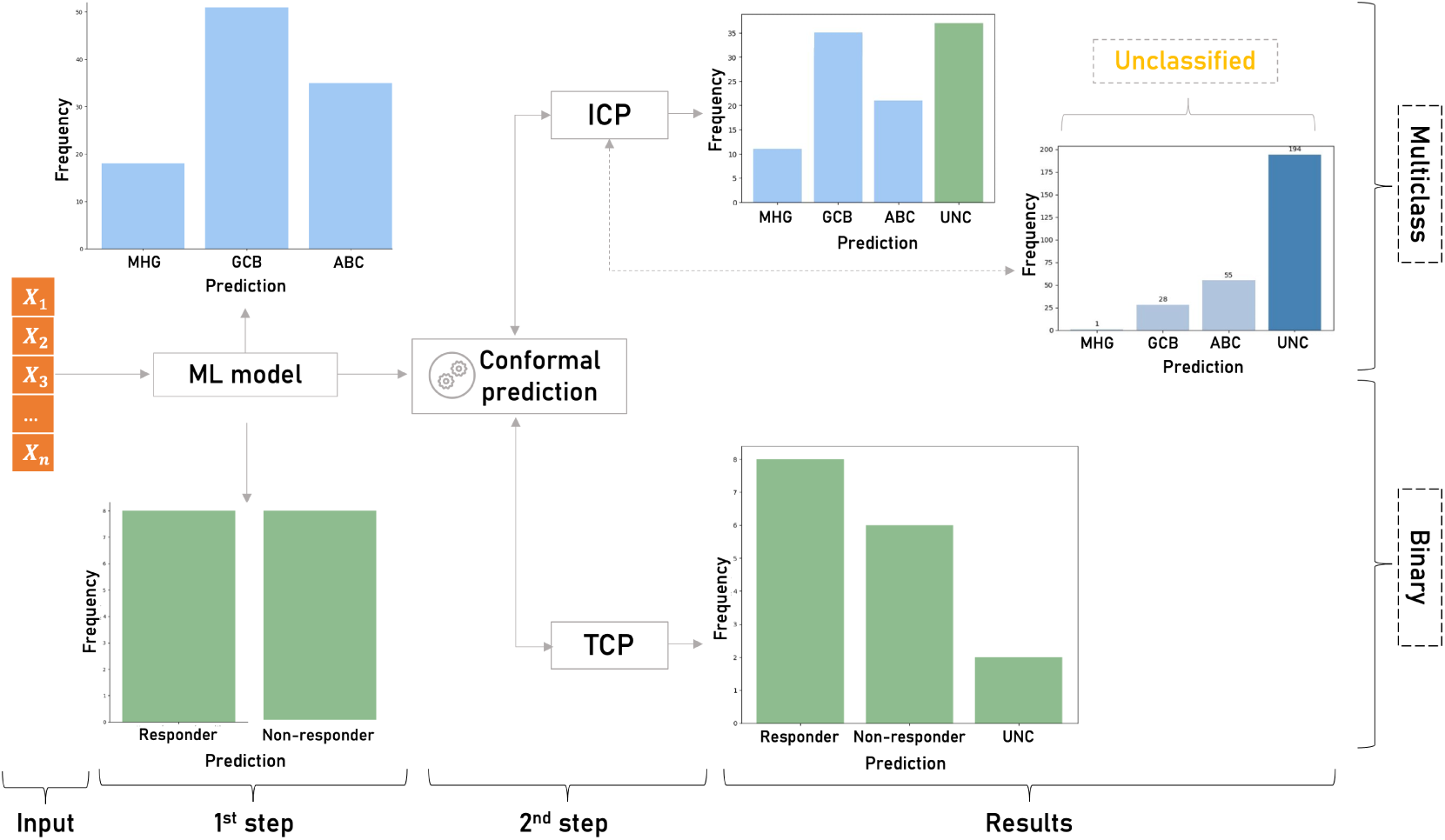
Overview of the classification study design. First, the ML model produces singleton predictions for the binary (responder/non-responder patients to infliximab) and the multiclass (MHG, GCB and ABC subtypes of diffuse large B-cell lymphomas) Singleton predictions are given without any indication of their accuracy and reliability. In the second step, CP is applied on the results of the ML models to estimate the uncertainty of each prediction. In the binary classification, TCP produces a prediction region that contains the true class with high probability and detects the uncertain (UNC) predictions. In the multiclass setting ICP identifies unreliable predictions among the samples classified by the nonconformalized model.

### 4.1 Responder prediction to infliximab

In their study, Mesko et al. correlated the pharmacological response of rheumatoid arthritis and Crohn’s disease patients to infliximab using their transcriptomic profiles. ^16^ The study includes 44 Crohn’s disease patients and 34 rheumatoid arthritis patients of which 40 responders and 38 non-responders. Affymetrix Human Gene 1.0 ST array quantified the expression levels of each sample in 33,297 target probes. The objective was to identify subsets of genes that can act as drug sensitivity biomarkers. In our experiment to sought to compare non-conformal and conformalized models in this binary setting using an ML model and a TCP framework to estimate the uncertainty of the model. TCP was selected as a favorable framework because it avoids the extra split for the calibration set which is preferable for small sample sizes. To evaluate TCP, we utilized the *empirical coverage* (Angelopoulos et.al, 2021), which measures the frequency of the true class within the prediction region. We then assessed the error rate threshold, ensuring it did not exceed the specified significance level of the conformal predictor.

Following the preprocessing step, we trained an SVM model on the top 100 genes with the highest discriminative power according to MRMR. For the 20% randomly selected patients included in the test set, the model yielded 87% accuracy (AUC=0.9), optimized by a grid-based parameter tuning (Fig. 2). By setting the significance level to 95% and defining the inverse probability, 1 *− p*(*y_i_|x_i_*), the probability of the model being incorrect, where *x_i_* is the feature vector and *y_i_* the label for the *i*th data point, as the *non-conformity measure* conformal predictions resulted in a 2.25% error rate compared to 12.5% of the SVM model without CP (Table 1). Two out of the 16 test cases were marked as uncertain requiring further evaluation by an expert physician. In this case, CP eliminated the misclassified samples by sorting out ambiguous cases, while for half of them the ML model alone made erroneous singleton predictions. The use of CP in this use case succeeded to reduce wrong predictions and to identify those cases that are hard to classify and should be forwarded for manual assessment.

**Table 1:**
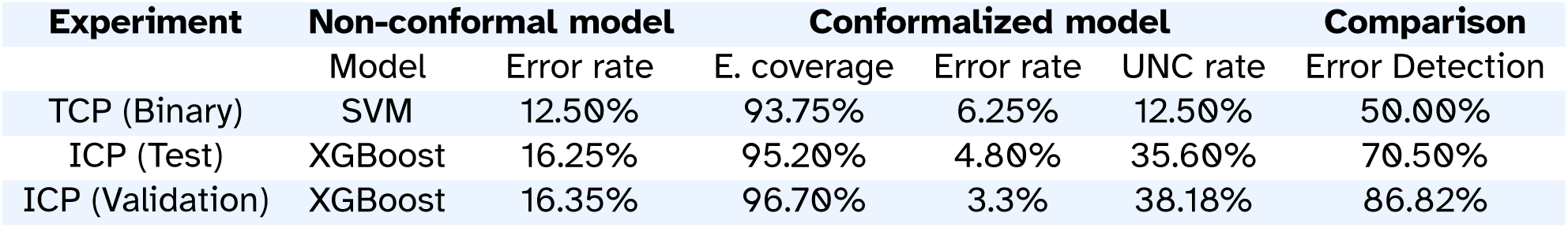
Performance of the non-conformal and the conformalized binary and multiclass models (95% confidence interval).

| Experiment | Non-conformal model |  | Conformalized model |  |  | Comparison |
| --- | --- | --- | --- | --- | --- | --- |
|  | Model | Error rate | E. coverage | Error rate | UNC rate | Error Detection |
| TCP (Binary) | SVM | 12.50% | 93.75% | 6.25% | 12.50% | 50.00% |
| ICP (Test) | XGBoost | 16.25% | 95.20% | 4.80% | 35.60% | 70.50% |
| ICP (Validation) | XGBoost | 16.35% | 96.70% | 3.3% | 38.18% | 86.82% |

Concerning the singleton predictions, TCP identified eight non-responders and six patients who will respond to infliximab. This group of patients is correctly classified to the actual class with an error rate of 5%. Moreover, for two wrong predictions of the non-conformalized SVM model, CP flagged one of them as uncertain, which identifies this patient as a difficult-to-classify case, (Table 1). For this patient, the treatment decision should be made by an expert. Overall in these personalized therapeutic decisions, CP can stand alongside the physicians to flag the difficult-to-predict patient cases for further manual data curation and closer treatment monitoring, thereby improving the decision-making time and minimizing the risk of wrong interventions.

### 4.2 Predicting molecular subtypes of diffuse large B-cell lymphoma

In the multi-class use case we used CP as a diagnostic predictor to classify patients with diffuse large B-cell lymphoma based on the distinct transcriptional profiles of their tumor cells. Diffuse large B-cell lymphoma is the most common hematological malignancy characterized by highly heterogeneous molecular signatures. Approximately 80% of the lymphomas are curable using R-CHOP combination therapy yet, there is a biologically heterogeneous group of patients that differs in terms of their clinical characteristics and prognostic factors. ^21^ Therefore to enable precise patient stratification in clinical trials, we first have to distinguish patients who are likely to respond to R-CHOP alone from patient groups who may benefit from emerging therapies based on the molecular heterogeneities of the disease.^17^

So far, diffuse large B-cell lymphoma patients are classified based on the Cell of Origin (COO) in the activated B-cell like type (ABC) and the germinal center B-cell like (GCB) subtypes. Recently, Sha et al. proposed a new distinct molecular subtype with aggressive clinical behavior called molecular high-grade B-cell lymphoma (MHG). ^18,19^ Patients of this subtype tend to not respond to R-CHOP therapy, despite the similarity with the GCB subtype, and they may benefit from either intensified chemotherapy or new targeted therapies. Clinical trials require the identification of the COO to personalize therapeutic interventions and to decipher the mechanisms of the disease pathogenesis.

In this experiment we built an inductive version of the CP model on a gene expression dataset of 1,311 samples extracted from formalin-fixed, paraffin-embedded biopsies (GEO Data series: GSE181063). The RNA samples include 345 ABC, 517 GCB, and 170 MHG molecular subtypes except for 278 patients who were not classified in any of the three classes and characterized as unclassified (Fig. 2). Illumina’s HumanHT-12 WG-DASL V4.0 beadchip array quantified the expression levels of each sample in 29,377 target probes. Following a data cleansing and quality control step 20 probes were selected by the MRMR algorithm to build the training feature set. The multi-class model was trained by XGboost, ^20^ the hinge loss function was applied as *nonconformity* measure in the ICP model and the empirical coverage was used to evaluate the conformal predictor.

XGBoost has a classification error of 16.25% on 10% of randomly selected patients. The conformalized XGBoost model resulted in 4.8% subtype classification error using 95% confidence level. In addition, the inductive predictor flagged, 37 patients (35.6%) as uncertain that are distributed in the following prediction regions: {MHG, GCB} = 8, {MHG, ABC} = 2, {GCB, ABC} = 19, {MHG, GCB, ABC} = 8. Non-singleton predictions involve mainly GCB samples that are most often misclassified as ABC samples. MHG has a clearly separable profile being transcriptionally closer to the GCB subtype. For eight patients the conformalized model was not able to exclude any prediction region. However, the ICP model managed to avoid the misclassification of 12 out of the 17 wrong predictions of the XGBoost model alone. The results reinforce the reliability of the prediction regions, as they detect the wrong assessments of the basic algorithm and give a better view of the difficult examples, while at the same time, they limit the range of possible classes to facilitate the final expert decision.

Concerning the 278 unclassified patients, although there is no class assignment ICP provides singleton predictions for 30.2% of the samples {MHG} = 1, {GCB} = 28, {ABC} = 55 (Fig. 2). The remaining are ambiguous cases involving either two classes {MHG, GCB} = 1, {MHG, ABC} = 9, {GCB, ABC} = 135, or all three {MHG, GCB, ABC} = 49. Among the 194 uncertain cases, most of them involve double predictions (145 cases) of GCB and APC classes, which is also inline with the principal component analysis in Fig. 3. Both single and double predictions provide insights beyond what a non-conformalized learning approach alone can offer and can be useful in preventing erroneous predictions that are of major importance in clinical decisionmaking.

**Figure 3:**
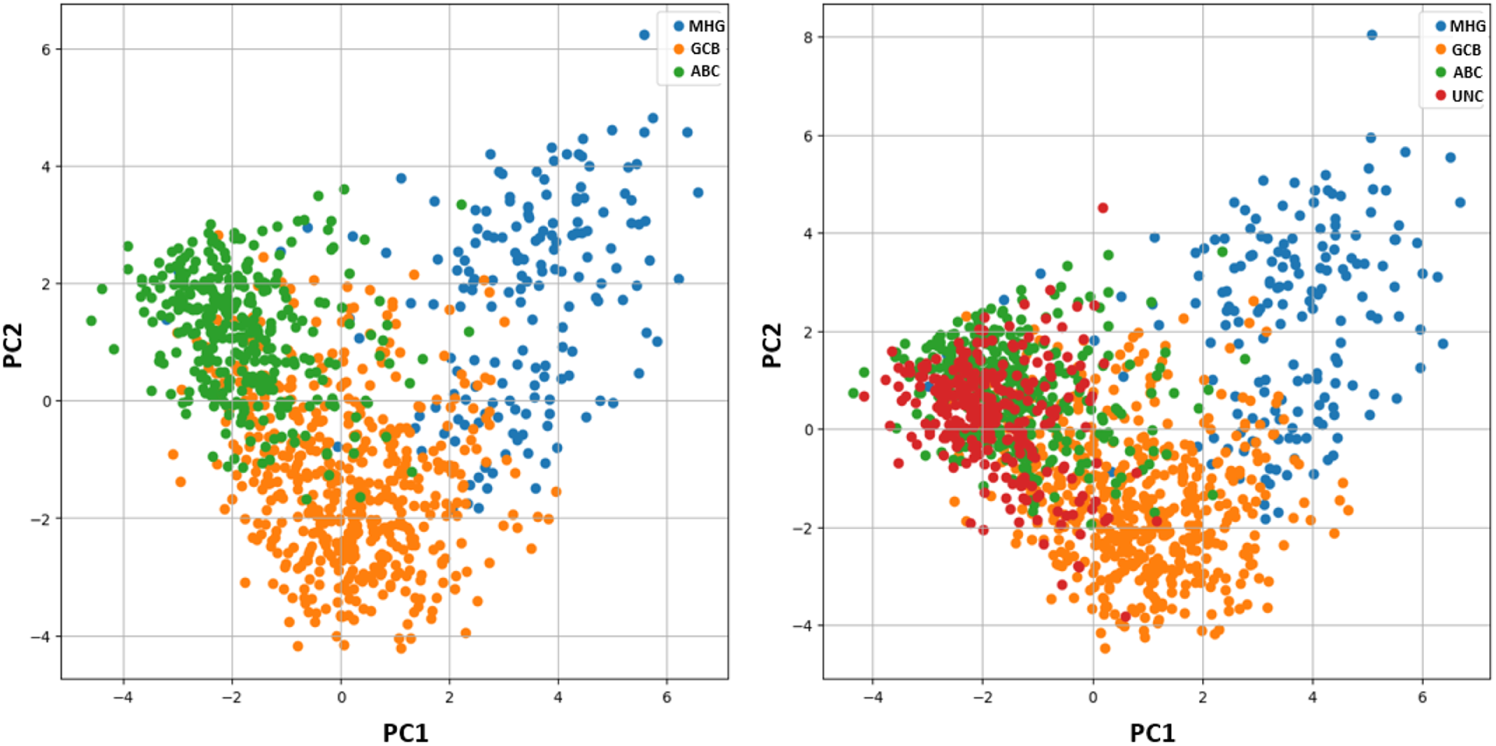
Principal Component Analysis (PCA). Left: PCA analysis of the samples without unclassified cases (UNC), revealing a small overlap between subtypes ABC and GCB. Right: PCA analysis of the dataset including unclassified cases (UNC), shows the UNC class overlapping with subtypes ABC and GCB.

To evaluate the reliability of the prediction regions on unseen data we sought to examine the fundamental exchangeability assumption on an external diffuse large B-cell lymphoma gene expression dataset (GEO Data series: GSE117556). The gene expression profiles were produced from 789 RNA samples extracted from formalin-fixed, paraffin-embedded biopsies using Illumina’s HumanHT-12 WG-DASL V4.0 beadchip array. To assess the level of distribution shift we applied the MMD measure and compared the produced probability distributions of the two datasets. Fig.4 shows the distribution shift between the two datasets. We computed an MMD statistic of 0.0011 and performed a permutation test to determine the p-value, which was found to be 0.017. Since this p-value is less than the significance level a=0.05, we reject the null hypothesis that the two datasets are generated from the same distribution. The robustness of the conformalized model under the distribution-shifted data was examined by estimating the classification performance of the ICP model on the external data. For a 95% significant level, the ICP model resulted in 26 misclassifications while the XGboost model alone failed to correctly classify 129 samples, out of the 789 samples. The ICP model flagged 112 out of the 129 misclassified samples as uncertain cases to be further assessed by clinical experts. The erroneous predictions of the conformalized model, limit by 80% the risk of failure on data under distributional shift. However, there is an increased number of double predictions that mainly involve the GCB and ABC samples accounting totally for 225 samples and 77 cases for triple predictions. On the contrary, the MHG class does not show significant overlap with the GCB and ABC samples, accounting totally for 2 and 14 cases, respectively.

**Figure 4:**
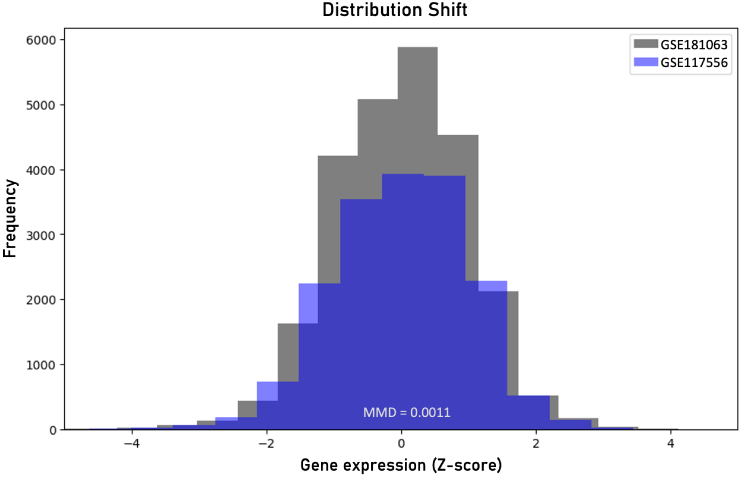
Visualization of the distribution shift between the two diffuse large B-cell lymphoma datasets based on the maximum mean discrepancy (MMD).

The results indicate that MHG has a separable transcriptional signature while deeper investigation is needed to accurately discriminate the GCB and ABC molecular profiles. Despite its ability to protect from false predictions, the conformalized model is more conservative than the XGboost model in making accurate singleton predictions (482 in total, of which {MHG} = 32, {GCB} = 295, {ABC} = 155). In other words CP effectively minimizes the risk of underestimating uncertainties at the expense of a lower number of definitive assessments.

Overall, in this generalizability test the ICP model achieved 96.6% empirical coverage on the unseen dataset. In principle, we proved the ability of our conformal classifier to generalize to data with different distributions, a significant advantage in medical applications where data heterogeneity is a common issue. These results, shown in Table 1, demonstrate the robustness of the conformal classifiers in handling data with varying distributions, but also the need to promote the adoption of CP-based frameworks in genomic medicine to be able to draw safer and more definitive conclusions.

### 4.3 Predicting pharmacological response of cancer cell lines to afatinib

In the regression use case, we implemented an ICP approach to predict the resistance of cancer cell lines to afatinib, an antineoplastic agent that is used to treat locally advanced and metastatic non-small cell lung cancer. Instead of categorizing samples into binary classes drug responses are quantified based on continuous drug concentrations that caused inhibition of 50% cell viability (IC50), with higher IC50 values indicating greater resistance. In this study, we sought to evaluate the scalability and robustness of CP uncertainty-aware regression models in predicting IC50 values using gene expression levels of cancer cell lines. The model was trained on a dataset of 765 cancer cell lines, each one including the expression levels of 17,613 genes and the corresponding IC50 values were recorded after 72 hours of afatinib treatment. The preprocessing steps, including outlier management and feature selection, refined the dataset to 677 cell lines with IC50 ranging between 0.00316 and 675, and identified 10 significant genes, meeting the i.i.d. assumption. A Random Forest (RF) algorithm was employed as the baseline regression model, the absolute deviation from the ground truth and the predicted value was used as non-conformity measure in the ICP model and the empirical coverage was used to evaluate the performance of the conformal predictor. The predicted IC50 values of the RF regressor had a mean squared error (MSE) of 14.35 and an R-squared value of 0.84 for 20% of the cell lines included in the test data.

As expected, the model exhibits significant deviation from the ground truth, mainly due to the heterogeneity of cancer types included in the dataset. To address this issue and better capture the variability in the data, we incorporated an inductive conformal predictor into the decision-making process. The ICP framework was employed to quantify the uncertainty of point predictions, providing a range within which the IC50 values are likely to fall. This approach aimed to reduce MSE and to provide a more precise and reliable estimation of the drug response for each cell line. Setting the significant level of 90%, and computing the non-conformity scores for the calibration set (20% of the training data), we found that at least 90% of the examples in the calibration set have a deviation value from the true IC50 value below the 6.77. The cutoff was set to 6.77 as it reflects the challenges faced by the baseline algorithm in accurately predicting new cases. With this value, we constructed the predicted range by adding and subtracting this value to every RF prediction. The conformal model constructed prediction ranges that contained the true IC50 value for 92.6% of the test set. When the same process was repeated with significance levels of 85% and 95%, the model achieved coverage rates of 90.6% for an a-Quantile of 6.63 and 95.6% for an a-Quantile of 6.98, as shown in Table 2. These results highlight the scalability of CP and its ability to meet user-defined coverage levels. Additionally, the a-Quantile in each case defined the size of the prediction intervals, with larger ranges corresponding to higher desired coverage levels and smaller ranges to lower ones.

**Table 2:** Performance of the ICP regression model.

| | Significance level | α-Quantile | E. coverage |
| --- | --- | --- | --- |
| 1 | 0.95 | 6.98 | 95.5% |
| 2 | 0.90 | 6.77 | 92.6% |
| 3 | 0.85 | 6.63 | 90.6% |

Overall, the regression conformal model effectively mitigated the inaccuracies of the baseline predictions by replacing individual point estimates with prediction intervals that achieve 92.6% coverage. This improvement is particularly significant in clinical settings, where constraining the index value to a high probability interval provides more actionable information than a single estimate with substantial potential deviation. Furthermore, instances where the true value falls outside the prediction interval serve as important indicators for further investigation, alerting experts for unusual cases that may require additional scrutiny.

## 5 Discussion

As the advancements in AI technologies are increasingly adopted into real-world problems the trustworthiness of ML applications in clinical environments is progressively acknowledged. However, denying taking a prediction risk when confronted with unusual cases is still not part of the mainstream procedures when building a model. CP is a powerful tool for estimating uncertainties as it combines favorable features such as i.i.d. assumption, the model-agnostic mode of application, and the adjustable prediction regions. CP addresses reliability concerns that often arise when dealing with imbalanced datasets, insufficient conditional coverage, and domain shifts. ^44^ Particularly in the genomics era, CP can overcome domain shifts caused by overlooking the heterogeneity introduced during data acquisition processes or data themselves, e.g. differences in the prevalence of a phenotype across populations. Coupled with larger or new representative calibration datasets under domain shift, CP provides adequate flexibility to keep coverage guarantees.

Another important feature is that CP can effectively lie on the top of both DL and ML models. The fundamental basis is that CP helps to quantify and communicate the model’s uncertainty effectively depending on the underlying model’s predictions. Traditional ML models typically deal with lower-dimensional features and simpler decision boundaries. These models typically provide clear decision rules or margins, which CP can straightforwardly translate into probabilistic measures of uncertainty. In contrast, DL operates on high-dimensional spaces with complex decision boundaries, capable of capturing intricate patterns and relationships in the data, which CP can use to generate more detailed and refined prediction intervals. The complexity of DL models allows CP to handle a wider range of applications in DL, such as image processing (Rouzrokh et.al, 2024), (Randl et.al, 2024), natural language processing**^? ?^** graphs or big data models.^49,59^ Thus, CP can adapt to the nature of the underlying model, utilizing the strengths of both traditional ML and DL to enhance the interpretability and trustworthiness of the predictions.

On the other hand, interpretability is another major concern, particularly in complex and blackbox deep learning models. CP has been acknowledged for its ability to provide guaranteed prediction sets and intervals that can be easily understood and communicated, offering a clear way to measure uncertainty. In addition, the minimal assumptions about the data distribution enhance interpretability by avoiding strict and probably unrealistic assumptions. However, the extent to which a conformalized prediction is interpretable partially depends on the interpretability of the underlying models themselves. For example, rule-based models, and decision trees offer a straightforward interpretation of their predictions contrary to deep neural networks and non-linear gradient boosting methods.

Although integrating conformal prediction into AI models seems compelling, there are a few limitations to be considered. Distribution-free uncertainty quantification methods, such as CP, are gaining interest among researchers due to their ability to provide reliable uncertainty estimates without assuming specific data distributions. CP ensures that, on average, can cover the correct class with a certain probability (marginal coverage). However, CP cannot provide guarantees for individual instances or structured subgroups of the data (conditional coverage).Practically, a conformalized model with a 90% marginal coverage guarantee ensures that the predictive set covers the correct class with 90% probability on average. This does not mean that each prediction covers the actual class with 90% probability for each individual instance or subgroup of the data. This limitation is crucial especially when dealing with specific subsets of data of particular interest, such as rare disease cases or minority classes in classification tasks. In such cases, researchers must be cautious when interpreting predictions, especially in scenarios where precise classification for individual instances is crucial.

In addition, challenges such as class imbalance, variance, and distribution shifts between training and validation data must be examined. These issues are mainly resolved by recalibrating the data using various combinations of attributes and classes with new data or by the adjustment of the existing calibration dataset with weights. Still, obtaining new and especially rare data to train the model with, in real-world scenarios can be challenging.

A reasonable question is how informative a conformal classifier can be in a binary classification setting when an uncertain prediction contains all the possible labeling options. Krstajic et al. question the utility of CP frameworks in binary classification scenarios.^39^ They reasonably wonder why someone should choose CP when a good binary classification model is built and how is it possible to include as correct coverage the predictions in which CP identifies both classes. In this work, we sought to highlight another aspect that is related to the detection of erroneous cases of the underlying model. In high-risk genomic medicine predictions, when specialists want to rely on the predictions of an ML model it is important to give them all the possible views of these predictions. For example, relying on a good binary classification model without any other guarantee of the resulting prediction may be a deterrent to incorporating such models into clinical decision-making. As we proved in the applications of this study, CP managed to detect the erroneous predictions of the underlying algorithm and classify them as uncertain cases. In clinical terms, these cases are translated as difficult to classify and consequently, the decision is risky to be taken by the ML model. In these cases, the contribution of an expert is necessary to avoid any misconduct.

In our point of view, the behavior of the conformal predictors can be a good step forward in bridging the trust between the medical community and the predictive modelling applications, since the latter can work side by side with the experts in the clinical decision-making process as a powerful and informative tool leaving the final decision to be deployed by experts in ambiguous cases. Additionally, a singleton prediction mathematically guarantees a safe decision with high confidence.

Overall, while conformal prediction offers valuable insights and uncertainty estimates for high-stake decision-making processes, it comes with several considerations and challenges. Ensuring the reliability of a prediction requires addressing issues such as distributional shifts in feature variables and labels, as well as the availability of representative calibration data. Although solutions such as recalibration and careful dataset management exist, they may not always be feasible, particularly in settings with limited data availability or where rare conditions are involved. Despite these challenges, the interpretability and robustness of conformal prediction make it a promising tool in domains such as healthcare, where the consequences of incorrect decisions can have a life-threatening impact and the ethical use of the models is mandatory. In-depth research and practical applications will be essential to address these challenges and to fully leverage the potential of conformal prediction in real-world scenarios. It is anticipated that as ML and genomic medicine are progressively infiltrating healthcare environments, CP will support more sophisticated approaches and enhance the range of uncertainty-informed multi-omics applications in clinical environments.

## 7 Authors’ contribution

C.P. implemented the methodology and wrote the manuscript, K.K. contributed to the design of the methodology, P.N. and I.C provided technical advice and reviewed the methodology, A.M. wrote the manuscript, conceived, and supervised the project. All authors reviewed the manuscript.

## 8 Data availability statement

The data and code used in this work are available at: https://github.com/BiolApps/ConformalPrediction

